# A Mixed-Effects Model to Predict COVID-19 Hospitalizations Using Wastewater Surveillance

**DOI:** 10.1101/2023.08.14.23293945

**Authors:** Maria L. Daza–Torres, J. Cricelio Montesinos-López, Heather N. Bischel, Colleen C. Naughton, Angel N. Desai, Marlene K. Wolfe, Alexandria B. Boehm, Miriam Nuño

## Abstract

During the COVID-19 pandemic, many countries and regions investigated the potential use of wastewater-based disease surveillance as an early warning system. Initially, methods were created to detect the presence of SARS-CoV-2 RNA in wastewater. Investigators have since conducted extensive studies to examine the link between viral concentration in wastewater and COVID-19 cases in areas served by sewage treatment plants over time. However, only a few reports have attempted to create predictive models for hospitalizations at county-level based on SARS-CoV-2 RNA concentrations in wastewater. This study implemented a linear mixed-effects model that observes the association between levels of virus in wastewater and county-level hospitalizations. The model was then utilized to predict short-term county-level hospitalization trends in 21 counties in California based on data from March 21, 2022, to May 21, 2023. The modeling framework proposed here permits repeated measurements as well as fixed and random effects. The model that assumed wastewater data as an input variable, instead of cases or test positivity rate, showed strong performance and successfully captured trends in hospitalizations. Additionally, the model allows for the prediction of SARS-CoV-2 hospitalizations two weeks ahead. Forecasts of COVID-19 hospitalizations could provide crucial information for hospitals to better allocate resources and prepare for potential surges in patient numbers.

## 1 Introduction

Wastewater-based disease surveillance (WDS) is a valuable tool for identifying and monitoring the spread of infectious diseases within a community. By detecting the presence of SARS-CoV-2 RNA in municipal sewer systems, WDS systems can provide early detection of outbreaks and track changes in the concentration of the virus over time. WDS can also be used to identify the spread of new variants when they emerge. Wastewater surveillance was used effectively prior to the COVID-19 (Coronavirus Disease 2019) pandemic, most notably as a tool to monitor and respond to poliovirus and to assess the presence of pharmaceutical and illicit drugs (Paul et al., 1940; Trask et al., 1938). The COVID-19 pandemic led to widespread implementation of WDS in communities across the United States. The approach has proven to be a cost-effective method for tracking COVID-19 and other infectious diseases (To et al., 2010; Hughes et al., 2022; Boehm et al., 2023a; Kumblathan et al., 2021). To bolster the nation’s capacity to monitor SARS-CoV-2 effectively, the US Centers for Disease Control and Prevention (CDC) launched the National Wastewater Surveillance System (NWSS) in September 2020, establishing over 1250 sampling sites across the US that encompassed more than 133 million people by October 2022. An essential factor contributing to the success of WDS is the extensive coverage of municipal wastewater collection systems, which connects approximately eighty percent of households in the United States.

In contrast, conducting widespread testing in a community requires substantial testing capacity, including access to testing kits, trained personnel, and testing facilities. Implementing community-wide testing can be financially burdensome, especially in resource-constrained settings. Even with adequate testing capacity, achieving high participation in testing can be challenging. When individuals get tested for a disease, their test-seeking and preventive behaviors can lead to biases. This can make the tested population less representative and affect the accuracy of disease prevalence estimates. Therefore, relying solely on confirmed cases to determine disease prevalence in a community can lead to biased estimates (Daza-Torres et al., 2023).

In order to reduce the biases introduced by testing practices, the test positivity rate (TPR) emerges as a crucial alternate measure. The TPR is the proportion of individuals who have tested positive for COVID-19 relative to the total number of tests administered. TPR has been used as a valuable alternative metric to track the spread of COVID-19 and assess the disease’s prevalence compared to the number of tests being conducted (Dallal et al., 2021; Dowdy and D’Souza, 2020). TPR has been demonstrated to reliably correlate with hospitalizations and intensive care units (Fenga and Gaspari, 2021; Montesinos-López et al., 2021). However, the interpretation and use of the TPR can be challenging in certain contexts, as it relies on accurate and comprehensive testing data, which may be hindered by factors such as limited testing capacity, variable testing practices, and variations in reporting standards.

One solution to the challenges faced in estimating disease prevalence in a community is through the use of WDS. This approach is relatively simple and accurate in gathering data from both symptomatic and asymptomatic individuals across an entire community. WDS provides a more accurate estimation of disease prevalence as it represents a sample of the population regardless of individual testing behaviors or access to healthcare. Numerous studies have observed a strong correlation between SARS-CoV-2 RNA concentrations in wastewater with the number of daily new cases in corresponding wastewater treatment plants catchment areas and, more recently, with the TPR (Montesinos-López et al., 2023; Boehm et al., 2023b).

Galani et al. (2022) and Kadonsky et al. (2023) offer examples of correlations between wastewater data and hospitalizations, with a time lag typically ranging from 8 to 14 days. Galani et al. (2022) implemented an artificial neural network model that combines clinical test results (COVID-19-positive cases) and wastewater concentrations to model new hospital and ICU admissions. These findings emphasize the potential of wastewater surveillance to improve preparedness and response to the spread of infectious diseases.

Building upon this knowledge, we propose using a linear mixed-effects model (LMM) to predict COVID-19 hospitalizations at a county-level by examining viral concentrations found in wastewater. Our analysis covers 21 counties in California for which wastewater data for COVID-19 is reported by WastewaterSCAN, a national WDS program that began in California in 2020 (Boehm et al., 2023d). The results of the present study thus offer an extensive assessment of correlations between wastewater data and hospitalization rates and demonstrate the broad applicability of the hospitalization forecasting models.

## 2 Materials and Methods

The analytical framework was developed using data from twenty-one counties across California, USA, which actively monitor wastewater. The analysis includes clinical and wastewater data from March 21, 2022, to May 21, 2023, covering three “waves” of the COVID-19 pandemic. We identified the waves by visually examining the data. We defined the first wave period from March 21, 2022 to October 15, 2022; the second wave starts on October 16, 2022 and ends on February 4, 2023; and the third wave starts on February 5, 2023 and ends at the end of the study period on May 21, 2023 (see Figure 1).

**Figure 1:**
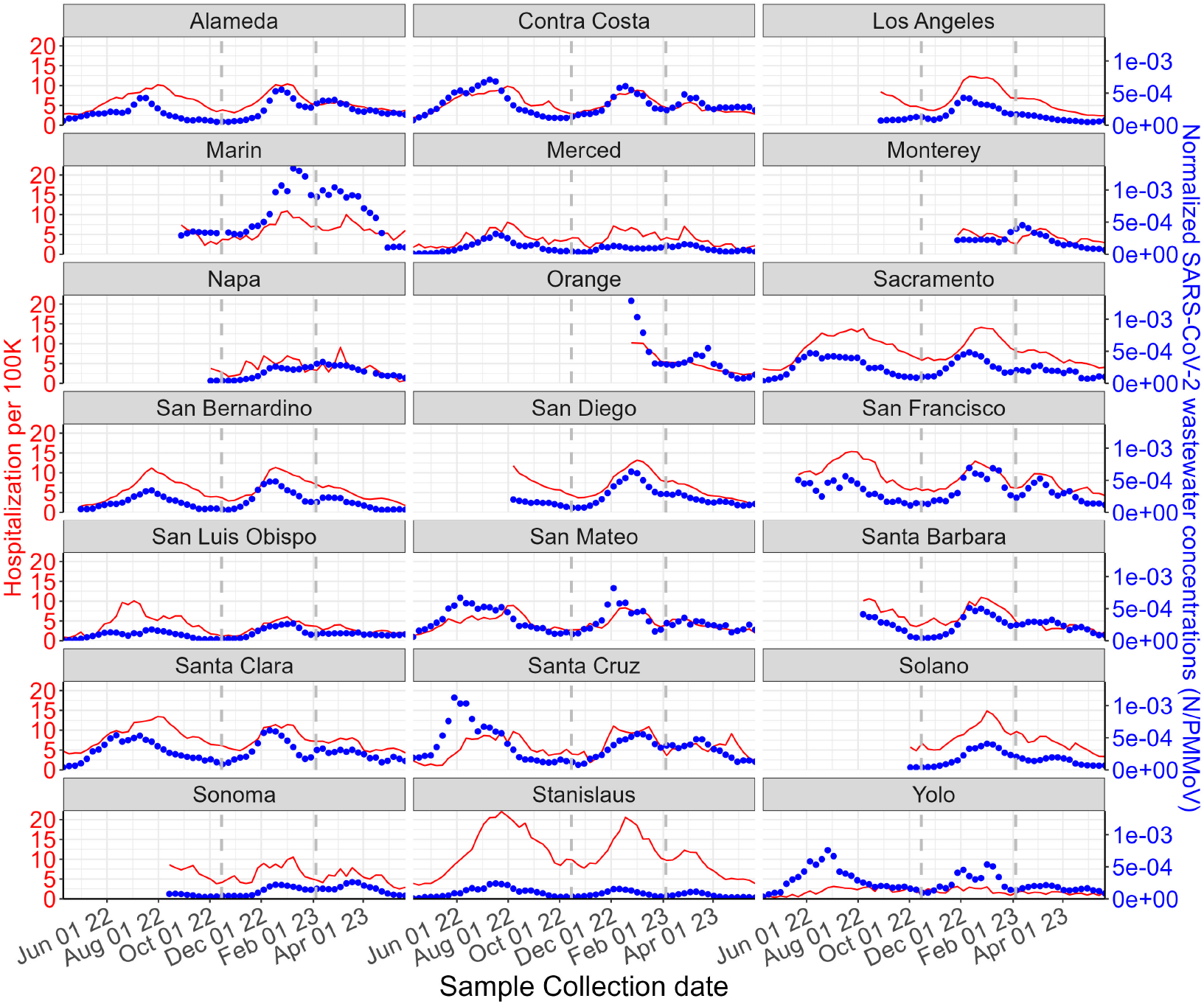
The weekly average COVID-19 hospitalizations per 100K population (red line) and weekly average normalized SARS-CoV-2 wastewater concentrations (N/PMMoV, blue dots) from March 21, 2022, to May 21, 2023, by county. Vertical gray lines represent the end of each wave.

### 2.1 Data Sources

#### Clinical Data

We used publicly available data for daily COVID-19 cases and hospitalization at the county level from the Official California State Government website, which provided the COVID-19 data from the California Health & Human Services Agency (CHHS). Hospitalization counts represent the number of patients hospitalized in an inpatient bed with a laboratory-confirmed COVID-19 diagnosis. It is important to note that some hospitalization data may involve individuals admitted to the hospital for reasons unrelated to COVID-19 but who tested positive during their hospital stay. The count includes all inpatients, including those in ICU and Medical/Surgical units, but excludes patients waiting for an inpatient bed in affiliated clinics, outpatient departments, emergency departments, and overflow locations.

The number of positive COVID-19 cases is determined by the total count of molecular tests that were positive using polymerase chain reaction (PCR) assays. We considered both “Total Tests” and “Positive Tests”, which reflect the cumulative totals based on the collection date. There is typically a delay between the date of specimen collection and when the test results are reported, which could impact real-time forecasts of hospitalizations as positive tests are reported.

Hospital admissions and COVID-19 cases were aggregated weekly. To calculate population-level hospitalization rates and cases for a county, we take the number of hospital admissions and new cases in the past 7 days, divide it by the population in that same county, and then multiply the result by 100,000. These metrics make it easier to compare data across different counties (refer to Figure 1). We will refer to the standardized hospitalization as “hospitalization rate” moving forward.

To calculate the TPR for a specific county, the number of positive COVID-19 tests from the past 7 days is divided by the total number of tests conducted in that county during that same time frame.

The analyses of this study used publicly available clinical data and do not contain any private health information. As a result, our study is exempt from ethical review and the requirement for informed consent for the clinical data, ac4cording to the Common Rule 45 CFR46.102 guidelines.

#### Wastewater Data

##### Sample collection

Samples were collected typically three times per week at 21 WWTPs as early as December 2021 and late May 2023 (Table S.1). Samples of settled solids were collected from the primary clarifier, or solids were obtained from raw influent by either using an Imhoff cone (Eaton et al., 2005), or allowing the influent to settle for 10-15 mins, and using a serological pipette to aspirate the settled solids into a falcon tube. Samples were collected by WWTP staff and sent at 4°C to our laboratory where they were processed immediately. The time between sample collection and receipt at the lab was typically between 1-3 days, during this time limited degradation of the RNA targets is expected (Guo et al., 2022; Burnet et al., 2023). Table S.1 provides additional information on the WWTPs including populations served and number and type of samples. In total, these WWTPs serve 13,893,677 people (approximately 35.7 percent of the CA population). A total of 4,764 samples were collected and analyzed.

The wastewater projects were reviewed by University of California, Davis. Written informed consent for participation was not required for this study in accordance with the national legislation and the institutional requirements.

##### Pre-analytical processing and RNA extraction

Detailed methods for pre-analytical processing and RNA extraction have been published in peer-reviewed papers (Wolfe et al., 2021; Daza-Torres et al., 2023; Boehm et al., 2023d) and on protocols.io (Topol et al., 2021a,b). In short, solids were dewatered using centrifugation, then one aliquot of solids was added to DNA/RNA shield (containing spiked in bovine coronavirus, BCoV, a process control) at a concentration of 75 mg/ml, a concentration at which inhibition of downstream analytical methods was minimized (Boehm et al., 2023d; Huisman et al., 2022). Another aliquot was used to determine the dry weight of the solids. The mixture was homogenized and centrifuged. Subsequently, nucleic-acids were extracted and purified from the supernatant using a commercial kit. The nucleic-acid extract was then processed through an inhibitor removal kit. The RNA was used immediately without storage as template in RT-PCR reactions, as explained below.

##### Analytical measurements

The RNA was used undiluted as template in digital droplet RT-PCR reaction containing previously described (Huisman et al., 2022) primers and probe for a target located in the N gene of SARS-CoV-2. This region has been conserved across all variants to date (Boehm et al., 2023b). The RNA was diluted 1:100 and used as template in a duplex digital droplet RT-PCR reaction containing previously published primers and probes (Wolfe et al., 2021) for pepper mild mottle virus (PMMoV) and BCoV. However, some specifics of the methods changed over time as the public health needs of the program changed requiring that various assays for other viral RNA targets were added and removed from the prospective monitoring program) and the number of samples processed by the lab increased, requiring a change in the number of replicates run for each sample (see Table S.2). These changes are described in Table S.2.

It should be noted that some of the data in this paper have been described in a Data Descriptor (Boehm et al., 2023d). Descriptions of the extraction and PCR negative and positive controls, BCoV recovery calculations, QA/QC elements, thresholding methods, and relevant Environmental Microbiology Minimal Information (EMMI) guideline reporting, are described in detail in the Data Descriptor. If BCoV recoveries were less than 10%, then the sample was rerun. The Data Descriptor describes data from San Jose, Southeast, SAC, and Davis collected through 12/31/22 and Merced and Modesto through 4/27/22 and 12/2/22-12/31/22. In addition, data from Merced and Modesto from 5/4/21 through 09/29/22 were included in a paper by (Kadonsky et al., 2023), and data from San Jose, Southeast, SAC, and Davis after 12/31/22 were included in a paper by Boehm et al. (2023c). The other data have not been published previously. All data used in this study are publicly available through the Stanford Digital Repository Boehm (2023).

The SARS-CoV-2 viral load is normalized by the PMMoV concentration, resulting in the dimensionless metric N/PMMoV. Previous mass-balance modeling work suggests that the ratio should be proportion to the number of SARS-CoV-2 RNA shedders in the contributing population (Wolfe et al., 2021). We calculated the weekly average N/PMMoV concentrations by dividing the sum of concentrations by the number of samples collected during that week. To analyze the relationship between hospitalization rates and SARS-CoV-2 RNA concentrations, we used data from the WWTP serving the highest population in each county. Figure 1 shows the N/PMMoV and hospitalization rate.

### 2.2 Data Analysis

This study focused on associations between hospitalization rates and longitudinal changes in the wastewater concentrations of SARS-CoV-2 RNA over time. Because COVID-19 hospitalization data were collected repeatedly through time for each county, the traditional linear regression model may not be appropriate due to the presence of correlated measurements within each county. Longitudinal studies often involve correlated data points for which the assumption of independence in linear regression is violated. LMMs allow us to explicitly model the dependence between observations within counties by incorporating random effects. We applied a logarithmic (base 10) transformation to the weekly average wastewater data and hospitalization rates to facilitate the analysis.

We hypothesized that the relationship between wastewater concentrations of SARS-CoV-2 RNA and hospitalization rate varies by county. This hypothesis is based on county-to-county differences in public health recommendations, public health access, and human behavior (Figure 2). We also aimed to consider the impact of time-dependent factors associated with each wave of infections. Time-dependent factors include virus variant evolution, human behavior changes, community vaccination status, and prior immunity. We accounted for these factors by incorporating a fixed effect into the model for discrete time periods associated with three waves of infections that were apparent over this study period (Figure 3). Kadonsky et al. (2023) observed that the wastewater-to-hospitalization ratios remained relatively stable during two COVID-19 waves in two geographically adjacent counties in California’s Central Valley between October 2021 and September 2022. This suggests that the wastewater-to-hospitalization ratios may remain constant in some cases, but the intercept may differ. Finally, we explore the predictive capabilities of various COVID-19 transmission indicators, including reported cases, TPR, and wastewater data. We aim to identify the most effective real-time indicator to monitor hospitalizations within a specific region.

**Figure 2:**
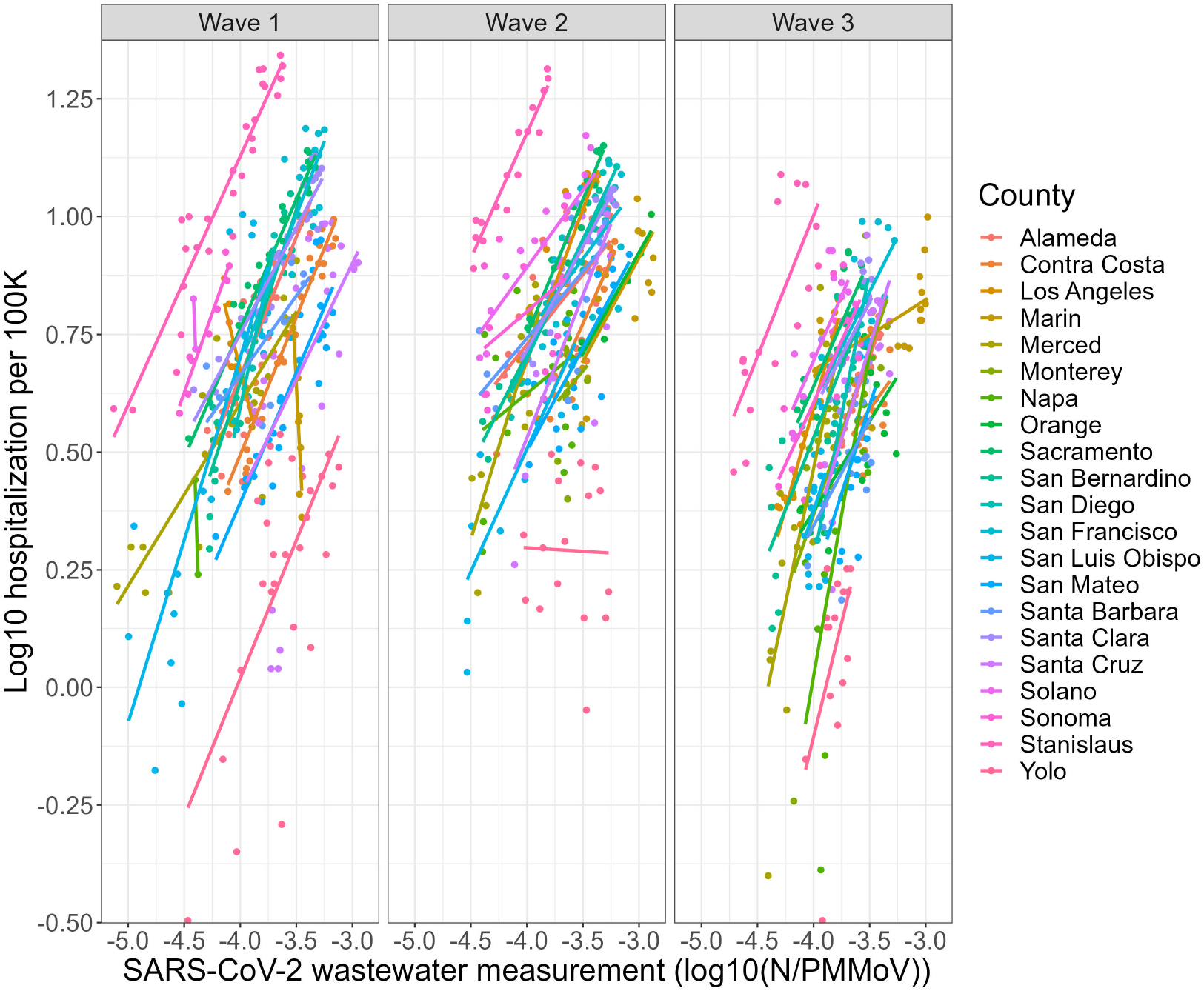
Log_10_-transformed weekly average COVID-19 hospitalization rate two weeks ahead of the wastewater sampling and log_10_ weekly average normalized SARS-COV-2 wastewater concentration (N/PMMoV) separated by the three waves of infection that were apparent over the study period. We identified the waves by visually examining the data. Regression lines are fitted to the data and depicted; model intercepts and slopes vary by county.

**Figure 3:**
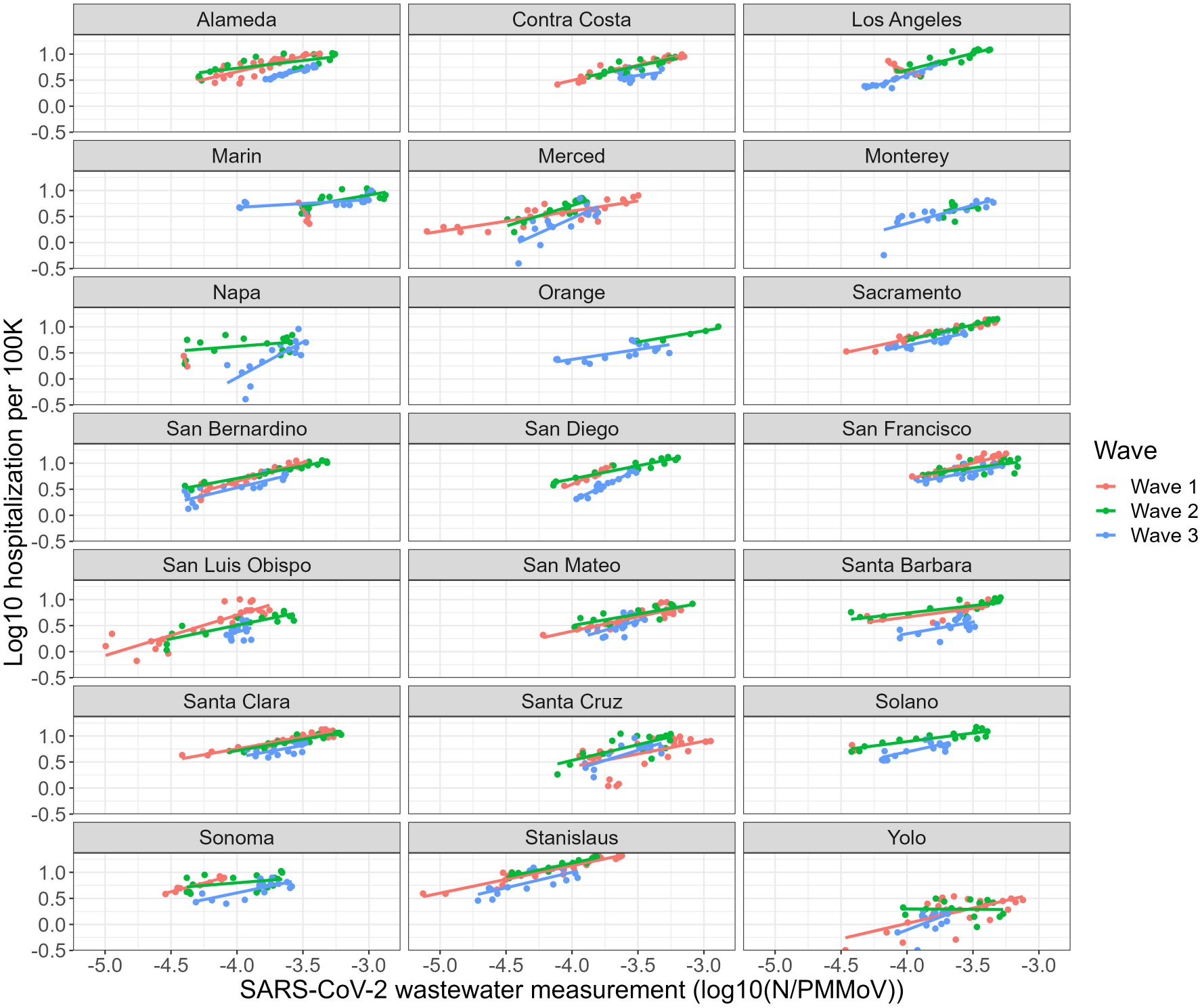
Log_10_-transformed weekly average COVID-19 hospitalization rate two weeks ahead of the wastewater sampling and log_10_ weekly average normalized SARS-COV-2 wastewater concentration (N/PMMoV) by county. Regression lines are fitted for each county and for each wave of infection considered.

### 2.3 Mathematical Model

The logarithmically transformed COVID-19 hospitalization rate at the county *i* = 1*, . . . ,* 21 and week *t* = 1*, . . . , T* is modelled as

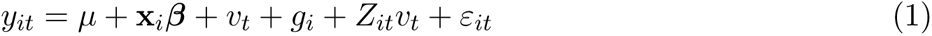

where *µ* is the average hospitalization rate across the study area and **x***_i_* is a vector of p covariates (e.g., wastewater concentration, reported cases, TPR; **x***_i_* = *{x*_1_*_it_, x*_2_*_it_, . . . x_pit_}*). The *v_t_*term corresponds to a temporal-level fixed effect indicating the pandemic’s three waves (grouped time component) as specified above. The *g_i_* term is a county-level random effect corresponding to the county where the hospital is located. County-level random effects are modeled as *g_i_∼ N* (0, *σ_2_*^2^), with *i* = 1, . . . , 21. The term *Z_it_v_t_* represents an interaction effect between a covariate *Z_it_* (e.g., wastewater concentration) and the wave factor. The *ε_it_* term represent the residuals distributed as *N* (0*, σ*^2^) with *σ*^2^ is the measurement error variance. The random effect *g_i_* is assumed to be independent of the residual *ε_it_*.

### 2.4 Model Implementation

The primary objective of this study is to evaluate our model’s ability to predict hospitalizations two weeks in advance using wastewater data. A two-week lag was incorporated to accurately capture the relationship between predictor variables and hospitalization outcomes. This lag duration was chosen based on previous research showing a nearly two-week delay between hospitalizations and measurements of wastewater, confirmed cases, and test positivity rate (Kadonsky et al., 2023; Montesinos-López et al., 2021). Specifically, we used the hospitalization data two weeks ahead (denoted as *Y_i_*) as the response variable for concentration, cases, and TPR at time *i*. While a single lag was selected in building the present model, the duration of the lag could be optimized for each county.

In order to account for the interdependence in hospitalization data, all models presented below will consider the variable “county” (i.e., the county where the hospital is situated) as a random effect in the predictor. The LMMs were executed using the “lme” function from the “nlme” package in R.

### 2.5 Model Evaluation

We tested 9 statistical prediction models to assess the model’s forecasting accuracy. We used the mean absolute scaled error (MASE) as our benchmark. To create these models, we used different predictors in Equation 1, including the number of confirmed cases (cases), TPR, N gene concentration (N), normalized N gene concentration (N/PMMoV), a wave factor (wave), and an interaction effects between normalized concentration and the wave factor (N/PMMoV*×*wave). The wave factor was added to account for several factors related to the emergence of a new variant, which cannot be measured directly.

The models M1, M2, M3, and M4 assume fixed effects predictor variables for cases, TPR, N gene concentration, and normalized N gene concentration, respectively. All four models also consider a county-level random effect. Models M5, M6, M7, and M8 add a wave effect factor to models M1, M2, M3, and M4, respectively. Model M9 is similar to model M8 but includes an interaction term between N/PMMoV and the wave factor. These models and their comparative metrics are summarized in Table 1.

**Table 1:**
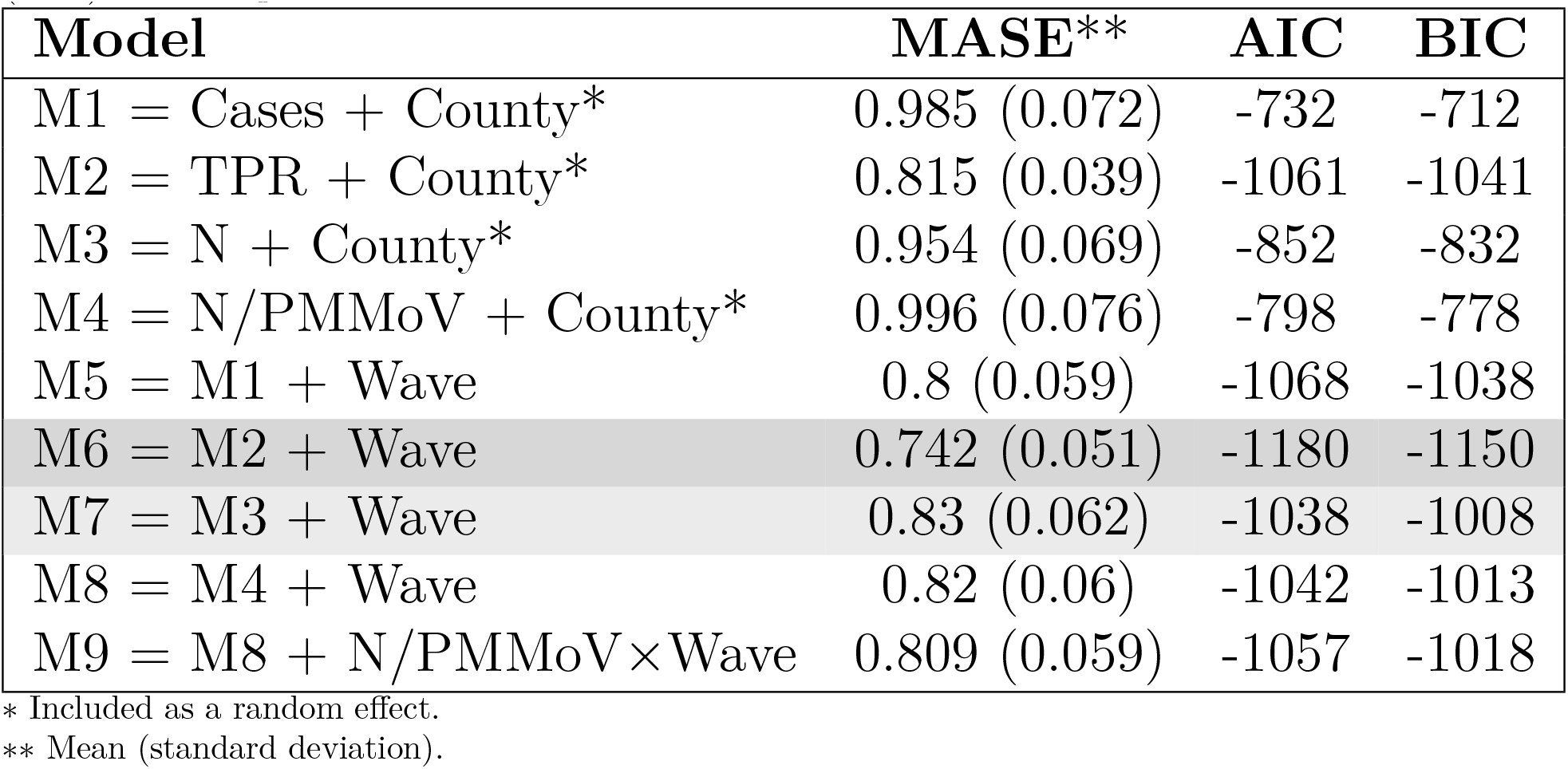
Model prediction performance based on the mean absolute scaled error (MASE), across 10 random partitions, assuming 80% of the total data set for training and the remaining 20% for testing. The Akaike Information Criteria (AIC) and Bayesian Information Criteria (BIC) are also presented.

To evaluate the performance of the models, we used ten-fold cross-validation. The data was split into training and testing sets at random. The training set received 80% of the data, while the testing set was allocated 20%. The model was created using the training set, and its prediction performance was evaluated using the test set. This process was repeated 10 times, creating 10 random partitions. The overall prediction performance was reported as the average of the 10 partitions.

The MASE was used to compare the observed values with the forecasted values to measure the accuracy of predictions. The MASE was computed across 10 partitions, 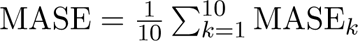, where MASE*_k_* of the k-th fold is estimated by where *y_t_* and *ŷ_t_* represent the observed and predicted values at time *t*, respectively. The Akaike Information Criteria (AIC) and Bayesian Information Criteria (BIC) were used to evaluate the goodness of fit, with lower values of AIC and BIC indicating superior model fit.

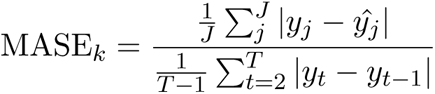

## 3 Results

Nine models were implemented to predict the number of COVID-19 hospitalizations in 21 counties across California (Table 1). We used the mean absolute scaled error (MASE) as a measure of the accuracy of the model forecasts. We calculated percent differences in MASE between corresponding models to highlight the effect of each covariate tested (Table 1 and Figure S.1). Models using TPR (M2 and M6) demonstrated the best performance overall. These models retroactively used the best measures of TPR, based on collection dates of clinical specimens. In real-time, measures of TPR are delayed by reporting times and may underperform when testing levels are overall low. Models that used wastewater data for hospitalization forecasts (models M3, M4, M7, M8, and M9) were considered successful for hospitalization forecasting when compared to models based on the traditional COVID-19 cases.

Adding a time-dependent factor for each wave of infections (models M5-M8) significantly enhanced forecasting accuracy compared to models without this factor (models M1-M4). Out of all these models, M5 showed the greatest improvement with the incorporation of a wave factor (18.8% better than M1), while M6 showed the least improvement with the addition of a wave factor (9% better than M2). Adding a wave effect to M3 and M4 improved performance by 13% and 17.7%, respectively. Normalization with PMMoV (model M8) did not improve prediction performance relative to the model that used unnormalized concentrations (model M7). We found that M2, based on TPR, outperformed models M1, M3, and M4 by 17.3%, 14.6%, and 18.2%, respectively. Models M1, M3, and M4 exhibited comparable prediction performance.

Model M9, which included an interaction effect between normalized concentration and the wave factor, did not improve compared to Model 8.

A cursory assessment of adding interaction terms to models M5 (casesxwave), M6 (TPRxwave), and M7 (Nxwave) indicated that these interaction terms also did not enhance model performance. The hospitalization rate predictions for California counties, based on Model M8, are presented in Figure 4. The predicted hospitalization rates align closely with the observed data, indicating that the forecasting model is performing well. Predictions for models M5, M6, and M7 are presented in Figs. S.2, S.3, and S.4, respectively.

**Figure 4:**
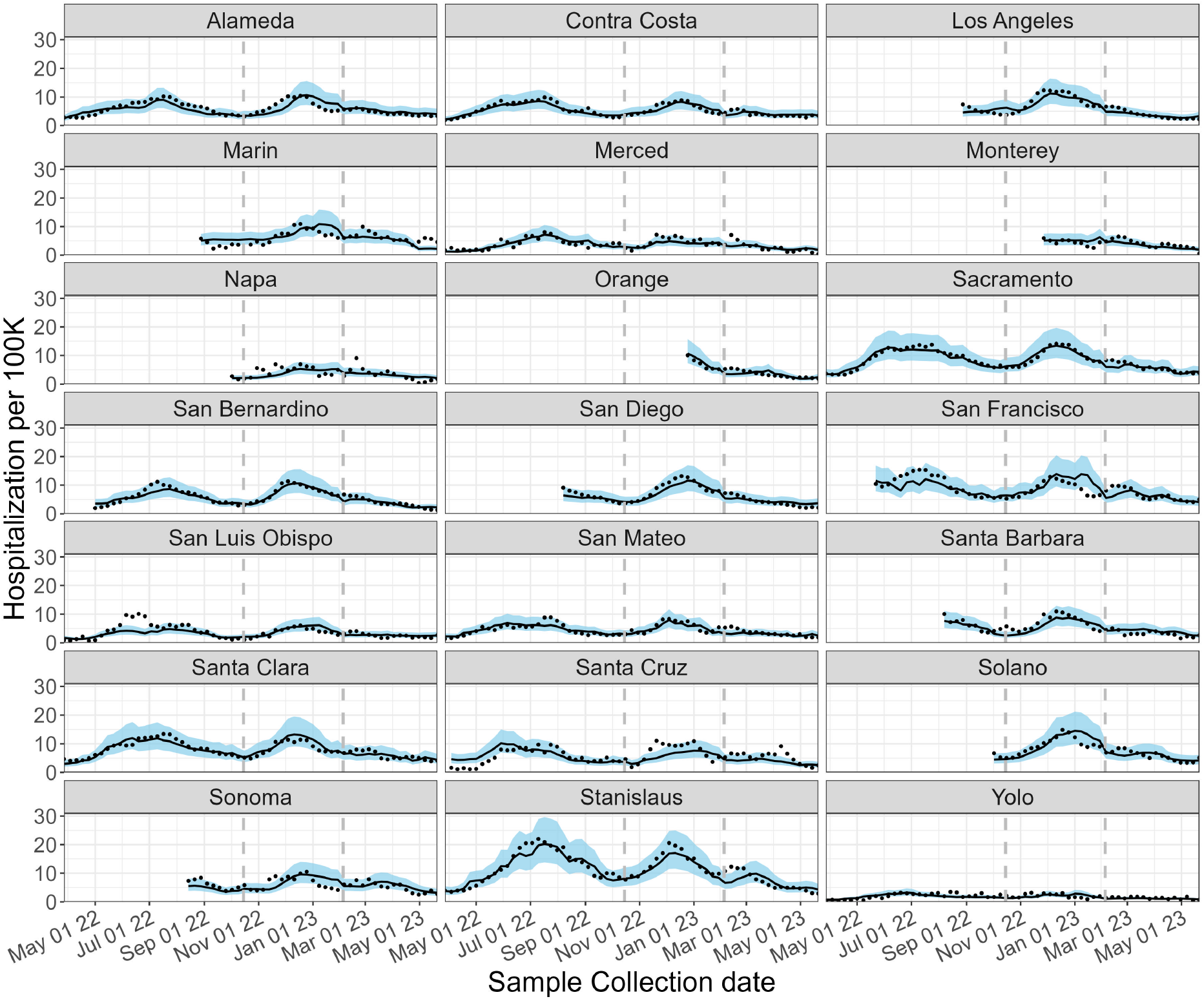
Estimated COVID-19 hospitalization rate based on Model 8. Black-solid lines and blue-shadow area describe the mean and 80% prediction intervals, respectively. Black dots represent the observed data. Vertical gray lines represent the end of each wave.

## 4 Discussion

Wastewater-based disease surveillance (WDS) models have been shown to be effective tools to estimate SAR-CoV-2 prevalence and effective reproductive numbers. WDS data has also been shown to correlate with hospitalizations. Our study demonstrates the feasibility of using WDS to reliably forecast hospitalizations at a county level, with a level of accuracy comparable to what we observed when using only confirmed cases or test positivity rates for hospitalization forecasting. The model forecasts yielded promising outcomes for WDS programs in 21 California counties, encompassing 13.5 million people across the state. Our findings thus offer a promising mechanism to improve real-time monitoring of infectious diseases using wastewater data and to support control measures for infectious diseases, especially when clinical testing is limited.

The mixed-effects models we developed facilitate evaluation of the associations between hospital admission rates for COVID-19 and three predictors: confirmed cases, TPR, and the concentrations of SARS-CoV-2 RNA in wastewater. We also introduced a time-dependent factor to account for the wave of infection underway. The wave factor recognizes the non-autonomous nature of the extended COVID-19 pandemic. The duration and spread of the disease through time are influenced by diverse elements including viral variant evolution, human behavior, vaccine development and uptake, prior immunity status, health interventions utilized, and other time-dependent variables. Inclusion of a wave factor enhanced the accuracy of all models tested. Model M6, which uses both TPR and the wave factor, yielded the strongest prediction performance amongst the nine models tested (lowest mean absolute scaled error). These findings echo previous research showing that TPR serves as a more reliable indicator of disease transmission than confirmed case counts (Dallal et al., 2021). Wastewater data yielded hospitalization forecasts with similar accuracy to models based on traditional testing data, indicating the significant potential for WDS programs to inform hospital capacity planning and resource allocation.

There are several limitations of our study. First, limitations of the hospitalization data must be recognized. Hospitalization data solely reflects the count of individuals who received a COVID-19 diagnosis while at the hospital. The hospitalization dataset thus includes individuals admitted to the hospital for reasons unrelated to COVID-19 but who tested positive during their hospital stay. It was not possible for us to disentangle hospitalizations that were not associated with COVID-19. Second, only 12 counties in the study had wastewater data for all three waves of infection that occurred during the study timeframe. As longer-term WDS datasets become available, future studies should continue to explore the effect of time-dependent factors associated with recurrent waves of infections. Third, our models did not incorporate interrelationships between communities or sewersheds (the areas covered by wastewater collection systems). We initially included a regional factor in the model, using regional classifications (Northern California, Greater Sacramento Region, San Joaquin Valley, San Francisco Bay Area, and Southern California). This broad geographic factor did not result in significant improvements in model performance and was eliminated in favor of model simplicity. More nuanced consideration of spatial correlation structures should be considered in future studies to assess the role of community interconnectivity and geographic proximity on disease dynamics. Finally, we recommend future studies incorporate an adaptive modeling framework. Adaptive frameworks that adjust model parameters according to new developments can generate more reliable and up-to-date forecasting to help decision-makers implement timely interventions.

Since the COVID-19 pandemic is no longer classified as a public health emergency, testing rates have diminished, and measures taken to control the spread of the disease have been curbed. Recent increases in COVID-19 cases and hospitalizations serve as a reminder to maintain vigilance. It is clear that WDS is a valuable tool for real-time disease monitoring, prevention, and preparedness. Using WDS to forecast hospitalizations offers an efficient method to translate early warning signals from wastewater into public health response strategies. By forecasting hospitalizations two weeks in advance, hospitals can be more proactive in managing their capacity to ensure that all patients receive the care they need. WDS infrastructure is also now being harnessed to monitor a suite of other infectious diseases. We believe there is tremendous potential to extend this approach toward other disease targets to inform hospital planning and public health decision-making.

## Data Availability

https://purl.stanford.edu/sh041sv7899
https://data.chhs.ca.gov/dataset/covid-19-hospital-data

https://purl.stanford.edu/sh041sv7899

## Availability Statement

All code and underlying data are publicly available through the GitHub repository (https:// github.com/Cricelio23/Hospitalization_WW). Analyses were carried out using R version 4.2.2.

## Acknowledgments

This research was conducted as part of Healthy Central Valley Together (HCVT), a collaborative research project to enable public health action from wastewater-based disease surveillance. Research support for HCVT was provided by a philanthropic gift to the University of California, Davis. The analyses conducted through Stanford University were supported by a gift from the CDC-Foundation and the Sergey Brin Family Foundation to ABB. We express gratitude to the staff at the wastewater treatment plants, public health entities, and others involved in the collection and curation of all data used in this research.

## Credit author statement

**M.L.D.T** and **J.C.M.L.**: Conceptualization, Methodology, Software, Formal analysis, Writing - Original draft preparation, Writing - Reviewing & Editing. **A.N.D**, **C.C.N.**, **H.N.B.**, **A.B.B**, and **M.K.W.**: Conceptualization, Methodology, Writing - Reviewing & Editing. **M.N.**: Conceptualization, Methodology, Supervision, Writing - Reviewing & Editing.

## Supplementary Material

**Table S.1:** Description of publicly owned treatment works (POTWs) and samples in this study.

| County | POTWs | Start date | N* | Population served | Sample type |
| --- | --- | --- | --- | --- | --- |
| Alameda | East Bay Municipal Utility District | 2/15/2022 | 199 | 740000 | Solids |
| Contra Costa | Central Contra Costa Sanitary District | 3/21/2022 | 167 | 484800 | Liquids |
| Los Angeles | Hyperion Water Reclamation Plant (HWRP) | 8/28/2022 | 114 | 4000000 | Liquids |
| Marin | Central Marin Sanitation Agency | 8/22/2022 | 71 | 104250 | Liquids |
| Merced | Merced Wastewater Treatment Plant (Merced) | 12/6/2021 | 218 | 91000 | Solids |
| Monterey | Monterey One Water - Regional Treatment Plant | 11/27/2022 | 69 | 262000 | Liquids |
| Napa | Soscol Water Recycling Facility | 9/26/2022 | 98 | 83300 | Solids |
| Orange | Regional Treatment Plant | 12/21/2022 | 64 | 129000 | Liquids |
| Sacramento | Sacramento Regional Wastewater Treatment Plant (SAC) | 12/6/2021 | 532 | 1480000 | Solids |
| San Bernardino | Regional Water Recycling Plant No.1 (RP-1) | 4/25/2022 | 164 | 890000 | Solids |
| San Diego | E.W. Blom Point Loma Wastewater Treatment Plant | 8/7/2022 | 120 | 2200000 | Liquids |
| San Francisco | Southeast San Francisco (SEP) | 5/20/2022 | 360 | 750000 | Solids |
| San Luis Obispo | City of Paso Robles Wastewater Treatment Plant | 1/11/2022 | 210 | 31037 | Solids |
| San Mateo | Palo Alto Regional Water Quality Control Plant | 12/6/2021 | 530 | 236000 | Solids |
| Santa Barbara | Lompoc Regional Wastewater Reclamation Plant | 8/1/2022 | 125 | 69290 | Liquids |
| Santa Clara | San Jose-Santa Clara Regional Wastewater Facility (SJ) | 12/6/2021 | 532 | 1500000 | Solids |
| Santa Cruz | City of Santa Cruz WTF - City Influent | 4/3/2022 | 172 | 160000 | Solids (Imhoff cone) |
| Solano | Fairfield-Suisun Sewer District | 9/28/2022 | 101 | 155000 | Solids |
| Sonoma | City of Santa Rosa, Laguna Treatment Plant | 8/11/2022 | 120 | 230000 | Solids (Imhoff cone) |
| Stanislaus | Modesto's Sutter Primary Treatment Facility (Modesto) | 12/6/2021 | 347 | 230000 | Solids |
| Yolo | City of Davis Wastewater Treatment Plant (Davis) | 12/6/2021 | 451 | 68000 | Solids |

\* N is the number of samples analyzed at each POTW.

**Table S.2:** Description of changes in the analytical measurement of the N gene in SARS-CoV-2 over time. The changes involved (1) the number of replicates from each sample from which nucleic-acids are extracted and subsequently run in individual digital droplet RT-PCR wells, and/or (2) how the N gene assay was multiplexed with other assays. Table columns include the names of the POTWs, and then each subsequent column shows the information on (1) and (2) for adjacent time periods indicated. S, XBB*, RSV, IAV, IBV, and HuNoV represent the S gene in SARS-CoV-2, 5 adjacent single nucleotide polymorphisms in the XBB* sublineages of SARS-CoV-2, respiratory syncytial virus, influenza A virus, influenza B virus, and human norovirus GII, respectively. BA.4 and BA.2 indicate assays for characteristic mutations in those SARS-CoV-2 variants (Boehm et al., 2023d).

| POTW(s) | Dates and method | Dates and method | Dates and method | Dates and method |
| --- | --- | --- | --- | --- |
| SAC, SEP, SJ | Start date through 3/12/23<br>Multiplexed as described in (Boehm et al., 2023d) with 10 replicates per sample | 3/13/22 - 5/21/23<br>Multiplexed with assays for XBB*, RSV, IAV, IBV, HuNoV using the QX600 6 color instrument from Bio-rad with 10 replicates per sample |  |  |
| Davis | 12/6/21 - 12/31/23<br>Multiplexed as described in (Boehm et al., 2023d) with 10 replicates per sample | 1/1/23 - 2/6/23<br>Multiplexed with IAV and S with 6 replicates per sample | 2/7/23 - 5/19/23<br>Multiplexed with IAV and XBB* with 6 replicates per sample |  |
| Modesto and Merced | Start date through 4/27/22<br>Multiplexed as described in (Boehm et al., 2023d) with 10 replicates per sample | 5/4/22 - 11/30/22<br>Multiplexed as described in (Kadonsky et al., 2023) with 3-5 replicates per sample | 12/2/22 - 2/6/23<br>Multiplexed with IAV and S with 6 replicates per sample. Modesto extra diluted ten fold prior to ddPCR as described in (Boehm et al., 2023d) | 2/7/23 - 5/19/23<br>Multiplexed with IAV and XBB* with 6 replicates per sample. Modesto extra diluted ten fold prior to ddPCR as described in (Boehm et al., 2023d) |
| All other plants | 2/15/22 (or other start date) through 5/26/22<br>Multiplexed with S and del143/145 with 6 replicates per sample | 5/26/22 - 6/14/22<br>Multiplexed with S and BA.4 with 6 replicates per sample | 6/15/22 - 7/11/22<br>Multiplexed with S and BA.2 with 6 replicates per sample | 7/12/22 - 2/6/23<br>Multiplexed with S and IAV with 6 replicates per sample<br>2/7/23 - 5/21/23<br>Multiplexed with IAV and XBB* with 6 replicates per sample |

**Figure S.1:**
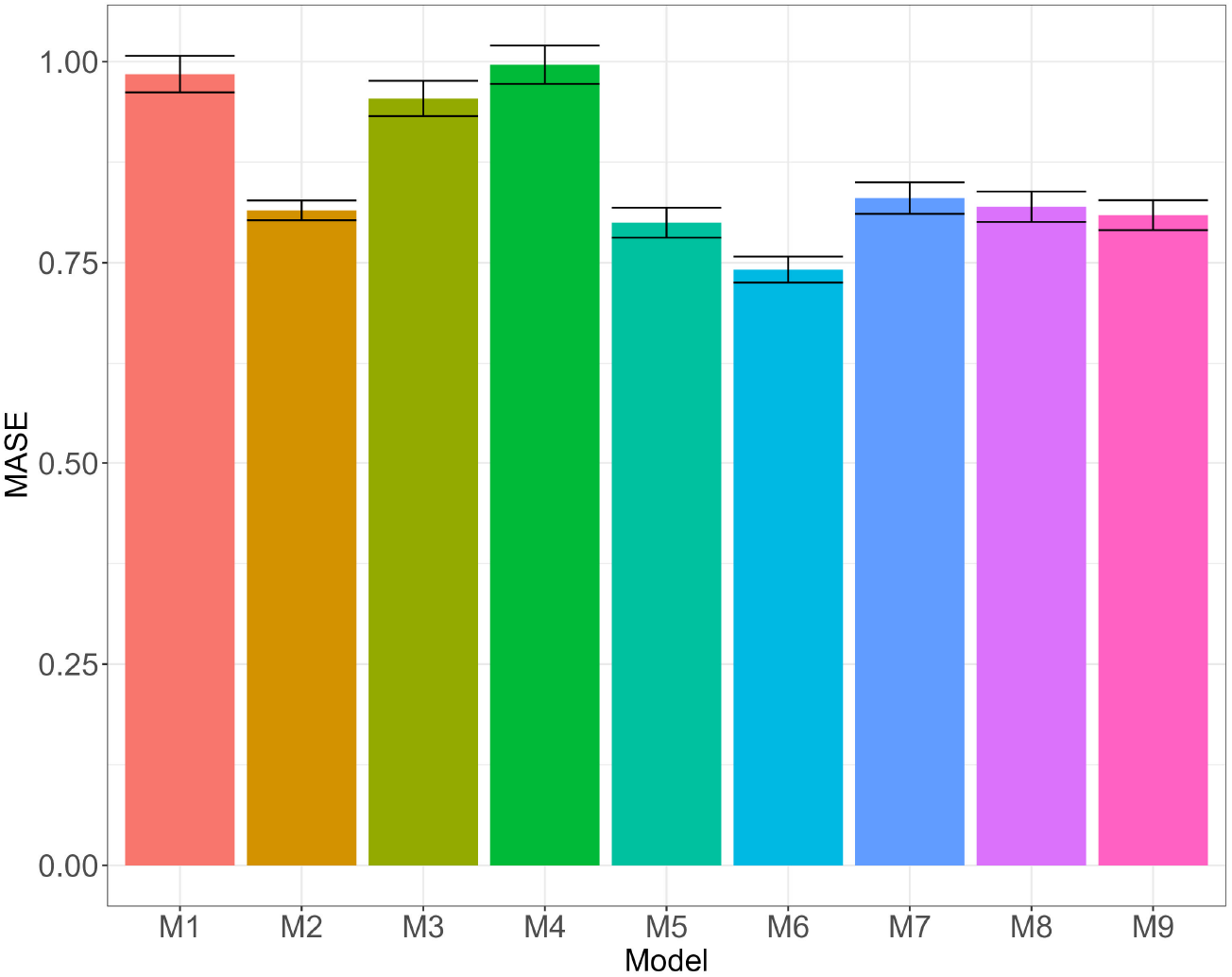
Performance prediction of models described in Table 1: mean absolute scaled error (MASE) with its standard deviation across the 10 partitions.

**Figure S.2:**
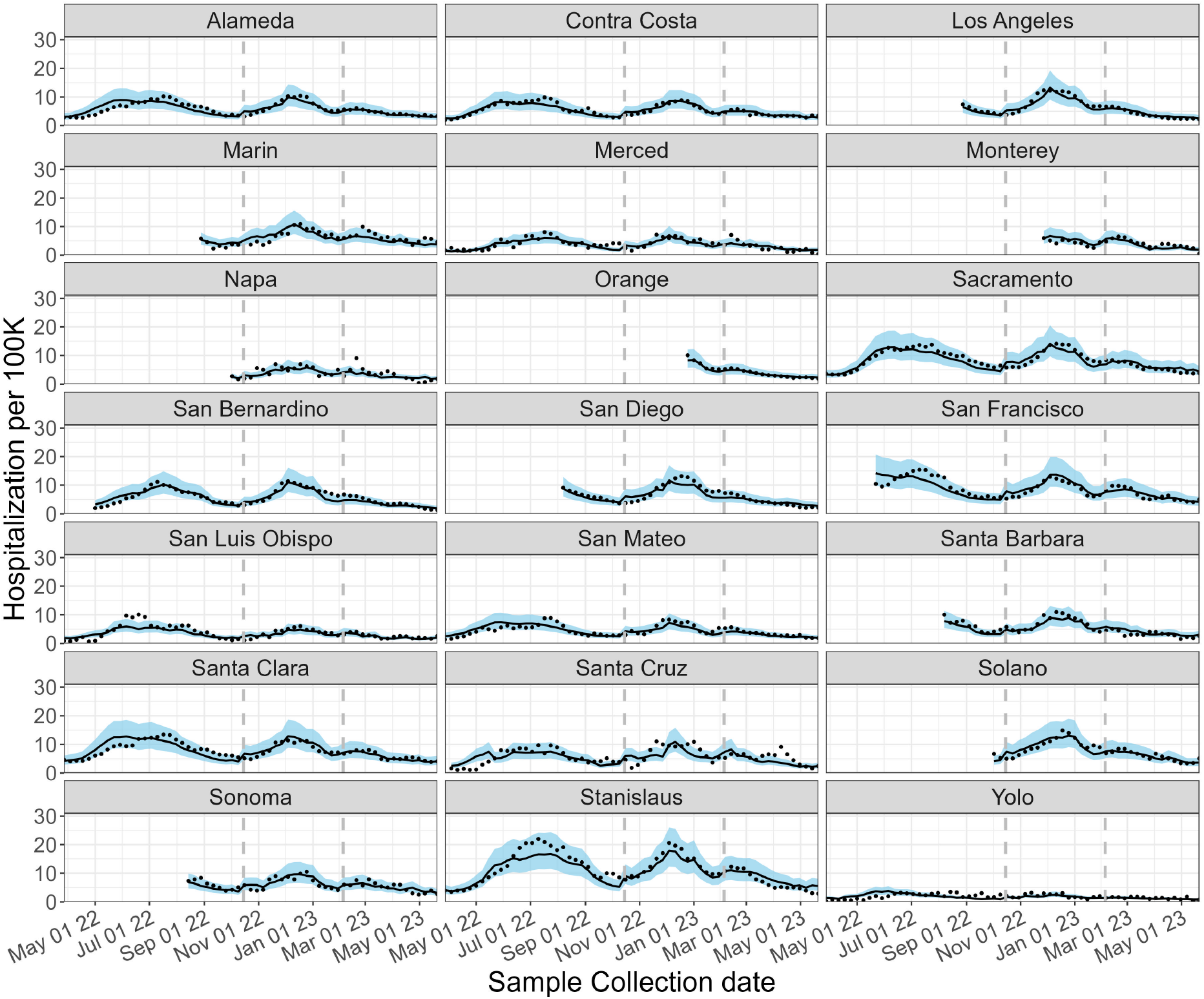
Model M5 = Cases + wave + County. Estimated COVID-19 hospitalization rate. Black-solid lines and blue-shadow area describe the mean and 80% prediction intervals, respectively. Black dots represent the observed data. Vertical gray lines represent the end of each wave.

**Figure S.3:**
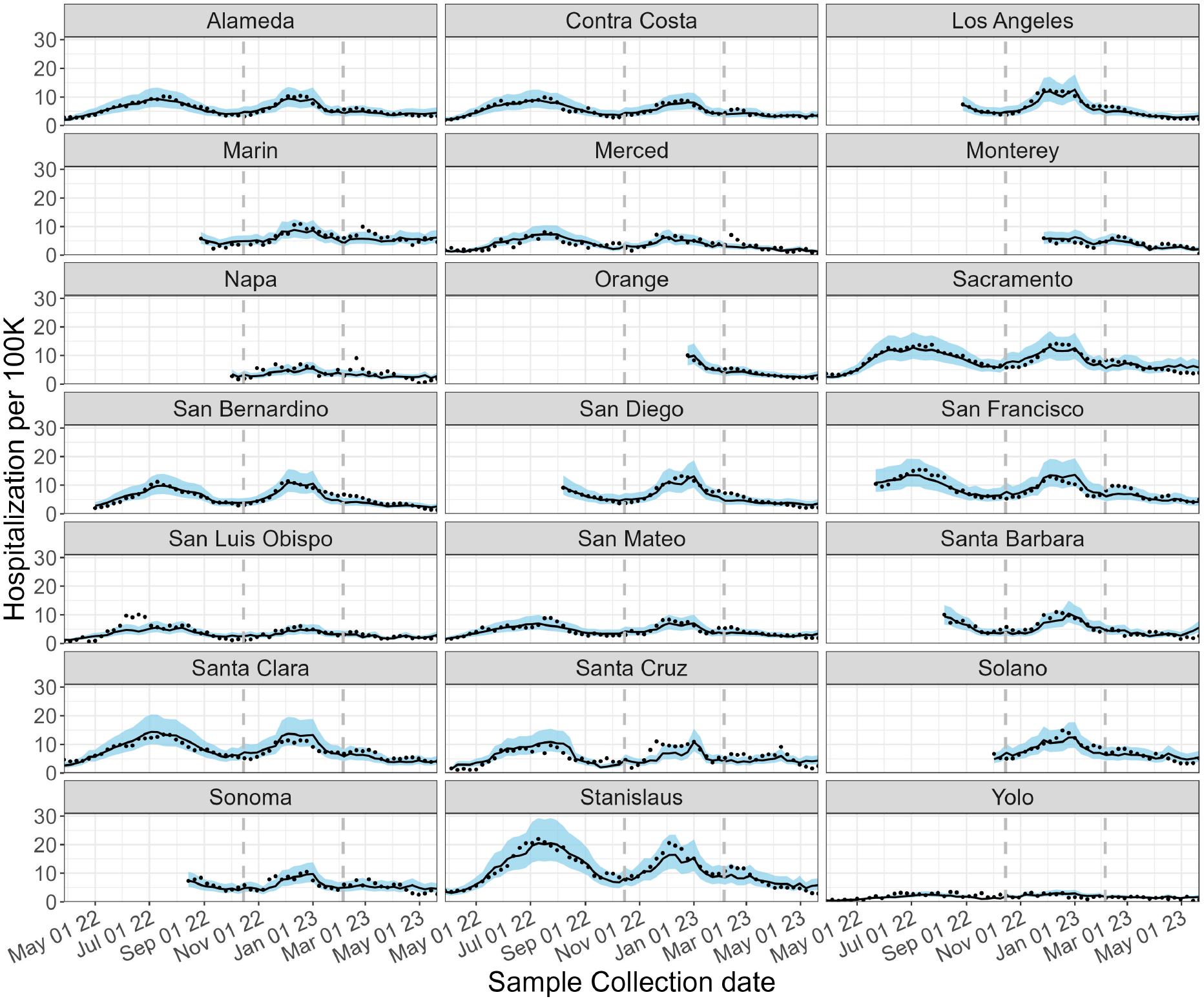
Model M6 = TPR + wave + County. Estimated COVID-19 hospitalization rate. Black-solid lines and blue-shadow area describe the mean and 80% prediction intervals, respectively. Black dots represent the observed data. Vertical gray lines represent the end of each wave.

**Figure S.4:**
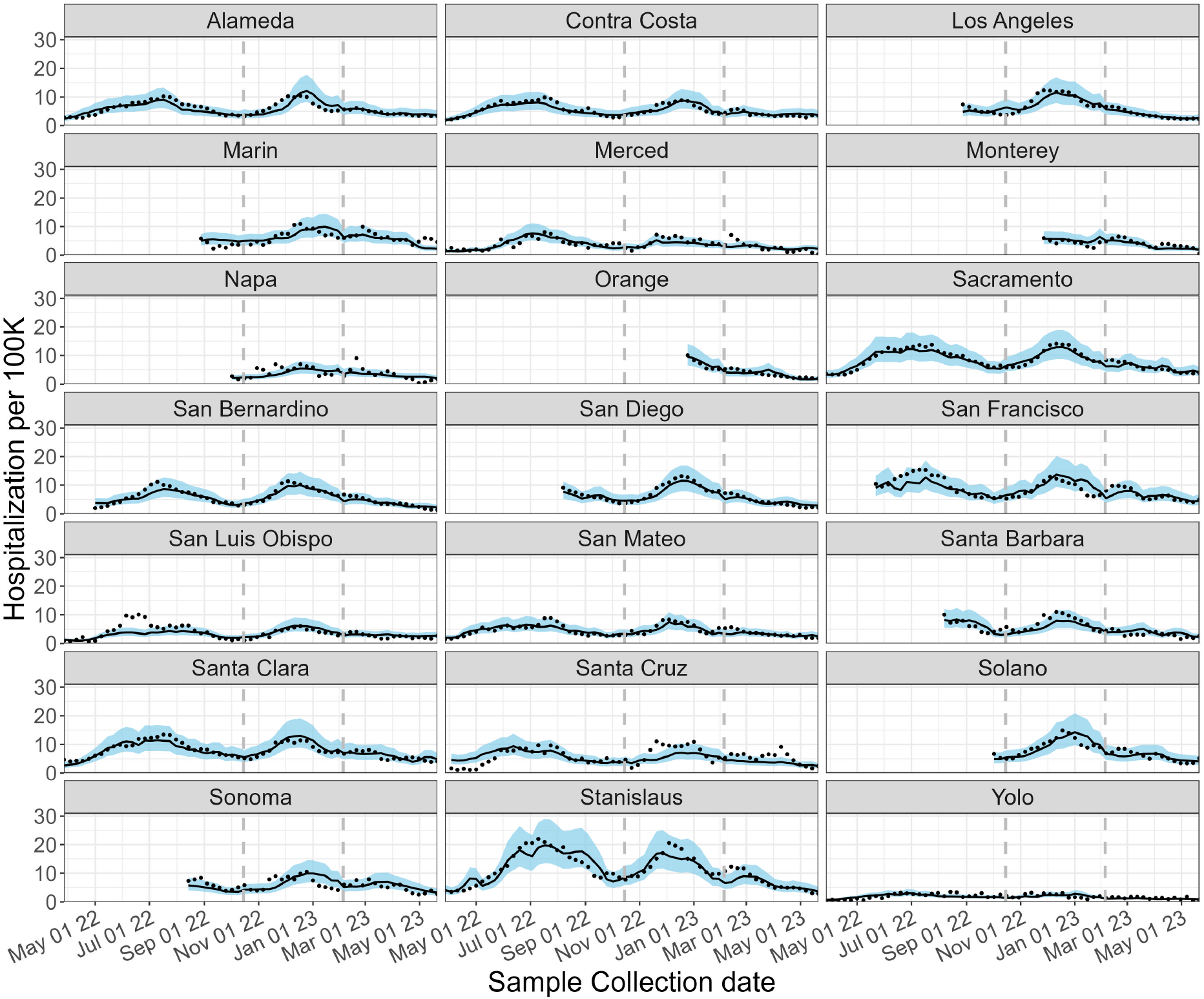
Model M7 = N + wave + County. Estimated COVID-19 hospitalization rate. Black-solid lines and blue-shadow area describe the mean and 80% prediction intervals, respectively. Black dots represent the observed data. Vertical gray lines represent the end of each wave.

